# Assessing the impact of testing for COVID-19 using lateral flow devices in NHS acute trusts in England

**DOI:** 10.1101/2024.06.06.24308561

**Authors:** Siyu Chen, Rachel Hounsell, Liberty Cantrell, Lok Hei Tsui, Reshania Naidoo, Prabin Dahal, Richard Creswell, Sumali Bajaj, Jennifer A. Flegg, Tom Fowler, Susan Hopkins, Ben Lambert, Merryn Voysey, Lisa J. White, EY-Oxford Health Analytics Consortium, Kasia Stepniewska, Rima Shretta

## Abstract

**Background:** Twice-weekly lateral flow device (LFD) testing was introduced for routine asymptomatic testing of healthcare workers (HCWs) in the National Health Service (NHS) in England in November 2020, with the primary aim of reducing nosocomial infections among staff and patients and a secondary aim of reducing absenteeism among HCWs. Here, we describe the burdens of HCW absenteeism and nosocomial infections in NHS acute trusts and the reported testing intensity of LFDs and associated costs from October 2020 to March 2022 and assess the impact of LFD testing on reducing these burdens.

**Methods and Findings:** We collected 16 million LFD testing results (total cost GBP 1.64 billion) reported in NHS acute trusts through England’s Pillar 1 and 2 testing programmes from 1 October 2020 to 30 March 2022. We estimated the prevalence of nosocomial COVID-19 infections in NHS acute trusts using data from the International Severe Acute Respiratory and emerging Infection Consortium (ISARIC). Testing data were linked with nosocomial infections and full-time equivalent (FTE) days lost by trust for NHS acute trusts.

We used a mixed-effects linear model to examine the association between FTE days lost and LFD test coverage. The relationship between weekly prevalence of nosocomial infections and LFD test coverage in the previous week was modelled using logistic regression weighted by the number of new COVID-19 cases reported in the ISARIC dataset for that week. We adjusted both models for community prevalence of COVID-19 infections, average income deprivation score, prevalence of variants of concern and LFD test positivity.

FTE days lost among HCWs varied considerably by trust type, staff group, geographical location of trusts, and progress of the pandemic in England. Increased LFD test coverage was associated with decreases in FTE days lost due to COVID-19 from November 2020 to July 2021, with no association observed from August 2021 to March 2022. Higher community prevalence levels were associated with significant increases in FTE days lost due to COVID-19 in all periods except the pre-vaccination period (last two months of 2020). The model predicted that changes in testing levels (50–150%) would have resulted in modest changes in FTE days lost due to COVID-19 for all time periods.

We identified 3,794 nosocomial infections (if patients developed COVID-19 symptoms 7 days or more after their hospital admission) among 106,377 hospitalised COVID-19 patients in 136 NHS acute trusts. The proportion of nosocomial infections among new weekly cases in hospitalised patients was negatively associated with reported LFD testing levels. The strength of the association varied over time and was estimated to be highest during the Omicron period, although no effect of testing on HCW absenteeism was found. The observed HCW testing/reporting was estimated to be associated with a 16.8% (95% confidence interval 8.2%, 18.8%) reduction in nosocomial infections compared with a hypothetical testing scenario at 25% of actual levels, translating to a cost saving per quality-adjusted life-year (QALY) gained of GBP 18,500–46,400.

**Conclusions:** LFD testing was an impactful public health intervention for reducing HCW absenteeism and nosocomial infections in NHS acute trusts and was cost effective in preventing nosocomial infections.

**Author Summary:** *Why was this study done?:*

- In any pandemic response, mass diagnostic testing plays a key role.
- We sought to evaluate the burdens of healthcare worker absenteeism and nosocomial infections in NHS acute trusts, the reported testing intensity using lateral flow devices (LFDs) and associated costs, and the impact of LFD testing on reducing these burdens.

*What did the researchers do and find?:*

- We collected 16 million LFD testing results and full-time equivalent (FTE) days lost due to COVID-19, obtained from healthcare workers (HCWs) in NHS acute trusts in England between 1 October 2020 and 30 March 2022.
- We estimated the number of nosocomial COVID-19 infections in NHS acute trusts using data from the International Severe Acute Respiratory and emerging Infection Consortium (ISARIC).
- Testing data were linked with nosocomial infections and FTE days lost due to COVID-19 by trust for NHS acute trusts.
- We used a mixed-effects linear model to examine the association between FTE days lost due to COVID-19 and LFD test coverage and applied a logistic regression to assess the association between nosocomial infections and LFD test coverage.
- We found that LFD testing in the healthcare setting was an impactful public health intervention.
- LFD testing reduced HCW absenteeism and nosocomial infections in NHS acute trusts; it was also cost effective in preventing nosocomial infections.

*What do these findings mean?:*

- Our analysis of the available data indicated that testing HCWs had varying impacts (on both nosocomial infections and HCW FTE days lost due to COVID-19) throughout the pandemic, possibly influenced by external factors such as community prevalence and vaccination.
- In any future pandemic, HCW testing interventions should incorporate collection of and/or timely access to relevant data, including HCW absenteeism, routine test results, community prevalence, and hospitalisation and mortality data.
- The lessons learnt from this study could be used by relevant authorities to support the real-time assessment of any testing service and adjustment of the testing regimen; they could also be used to help develop more targeted and agile testing systems, which operationally would require the ability to turn mass testing off and on as an epidemic progressed.

## Introduction

The COVID-19 pandemic led to a huge increase in the number of people requiring medical care, putting immense pressure on healthcare workers (HCWs) and highlighting the importance of their resilience and well-being. In England, SARS-CoV-2 infection rates among HCWs were estimated to be at their highest between April and September 2020 and decreased progressively over time until October 2021 [1]. HCWs were seven times more likely to experience severe COVID-19 infection than individuals with other, ‘non-essential’ jobs during the first UK-wide lockdown [2]. This was due to HCWs’ higher frequency of contact with suspected or confirmed COVID-19 cases, leading to a substantial burden of HCW absenteeism. Infected HCWs could inadvertently transmit the virus to vulnerable patients, exacerbating the spread of the disease; during the early stages of the pandemic, nosocomial infections were estimated to be responsible for 20% of all identified COVID-19 cases in hospitalised patients [3]. Regular testing for COVID-19 helped identify asymptomatic or pre-symptomatic cases among the healthcare workforce, allowing for prompt isolation and reducing the risk of transmission within healthcare settings [4].

Table 1 provides a summary of the time periods of the different stages of the COVID-19 pandemic and their associated testing policies and epidemiological features. Lateral flow device (LFD) testing was introduced for routine asymptomatic testing of HCWs in England in November 2020; HCWs were requested to undertake twice-weekly LFD testing at home with a follow-up confirmatory polymerase chain reaction (PCR) test in the case of a positive LFD result [5]. This large-scale testing campaign was initially conducted by NHS Trace and Trace, and later by the UK Health Security Agency (UKHSA), with the intention to support the NHS in its infection control risk reduction strategy and reduce staff absenteeism [6]. HCWs with confirmed COVID-19 infections were initially required to self-isolate for 10 days [7], which was the same as the self-isolation guidance for the general population [8]. Testing policy changed over time in response to the evolving epidemic and the vaccination campaign. The length of isolation was reduced from 10 to 7 days in December 2021 in the event of a negative LFD test result, with a further reduction to 5 days in January 2022 [9]. Confirmatory PCR tests following a positive LFD test result were no longer required from January 2022 [10].

**Table 1.**
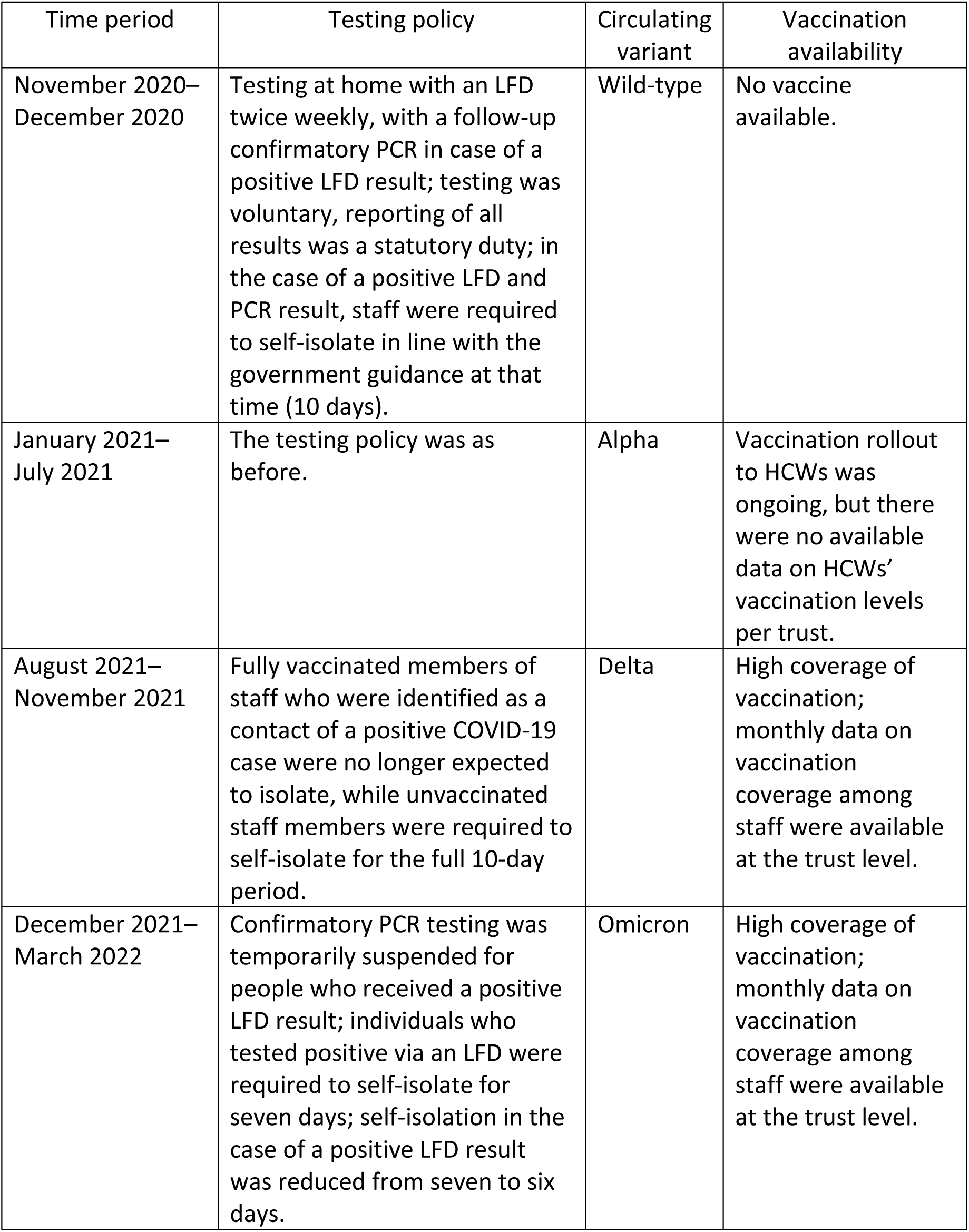
Summary of time periods during the COVID-19 pandemic and their associated testing policies and epidemiological features.

Here, we sought to determine the burden of absenteeism and nosocomial infections due to COVID-19 among HCWS in England between November 2020 and March 2022. We also explored correlations between these burdens and LFD test coverage in National Health Service (NHS) acute trusts in England and the associated costs.

## Materials and Methods

### Data acquisition and sources

#### LFD test volume and costs

LFD test volume data obtained via England’s Pillar 1 and Pillar 2 testing programmes [11] from 1 October 2020 to 30 March 2022 in healthcare settings were provided by UKHSA, in the context of a retrospective evaluation of England’s COVID-19 testing policy [12, 13]. Only data from NHS acute trusts were identifiable in both datasets, so only these data from the healthcare sector were analysed. LFD test costs during the evaluation period were extracted from internal UKHSA documents and derived from the Cost Allocation Project (an unpublished internal project) conducted by UKHSA for the UK’s Office for National Statistics (ONS); an analysis of these costs was performed as part of the retrospective evaluation [13].

#### NHS trust headcount and full-time-equivalent days lost

Monthly numbers of NHS Hospital and Community Health Service (HCHS) staff working in NHS trusts in England, presented as headcount and full-time equivalent (FTE) figures from 1 October 2020 to 30 March 2022, were retrieved from NHS Digital [14]. FTE days lost due to COVID-19, defined as the proportion of FTE days available that were lost due to COVID-19, were also downloaded from NHS Digital. As test data at the trust level were not available by staff groups, and as there were no specific recommendations throughout the evaluation period regarding which staff groups should be testing, we conducted the analysis with reference to FTE days lost due to COVID-19 for all staff.

#### COVID-19 community prevalence in England

We calculated weekly estimates of COVID-19 community prevalence for each lower-tier local authority (LTLA) within which each NHS acute trust was situated. These estimates were generated using a causal debiasing methodology [15], which used high-quality, randomised surveillance measures of swab-positive results provided via the REACT-1 survey [16] to debias PCR testing data obtained through Pillar 2.

#### Variants of concern (VOCs)

Weekly counts of SARS-CoV-2 lineages by week and LTLA were obtained from the Sanger Institute website [17]. These counts were then binned according to whether they corresponded to the Alpha (B.1.1.7), Delta (B.1.617.2) or Omicron (B.1.1.529) variant or whether they were from ‘other’ lineages. These data were then used to determine weekly LTLA-level proportions of each of these VOCs.

#### Healthcare worker vaccination

Data held in the NHS Electronic Staff Record (ESR) regarding the numbers of NHS acute trust HCWs who had been vaccinated for COVID-19 were retrieved from the NHS National Immunisation Management System (NIMS) [18]. Cumulative vaccination data for HCWs were available from the end of August 2021, when high coverage for the first (85%) and second dose (75%) had already been achieved.

#### Average income deprivation scores

The English indices of deprivation provide information on the prevalence of multiple types of deprivation in England at the LTLA level. We extracted these data from the UK ONS on 1 November 2022. LFD test data, community COVID-19 prevalence and average income deprivation scores were provided per LTLA rather than per acute trust. Therefore, we used a function from the covid19.nhs.data package [19] to calculate the number of tests per NHS acute trust in England, based on the proportional contribution of the number of tests conducted in LTLA regions within trusts (henceforth we refer to this as the ‘mapping function’).

#### ISARIC (International Severe Acute Respiratory and emerging Infection Consortium) database

We requested the relevant dataset from the ISARIC Data Access Committee. Weekly numbers of new COVID-19 admissions or hospitalised patients newly diagnosed with COVID-19 from 136 NHS acute trusts were extracted. Among these cases, COVID-19 infections were classified as nosocomial infections if patients developed COVID-19 symptoms seven days or more following their hospital admission. For a sensitivity analysis, we repeated these analyses using a 14-day cut-off. We included in our analysis all patients who were tested, had confirmed COVID-19 infection and were treated in an NHS acute trust in England. We excluded any re-admissions, records with conflicting dates, admissions outside of the study period (date of admission later than 31 March 2022 or date of discharge earlier than 1 October 2020), and COVID-19 cases outside of the study period (date of onset before 1 October 2020 or after 14 April 2022). For each week and for each acute trust in the ISARIC database (after exclusions), the total number of new COVID-19 infections, defined as new COVID-19 admissions or hospitalised patients with new COVID-19 symptoms, was calculated.

#### Hospitalisations and deaths data

Daily new hospitalised cases of COVID-19 in acute trusts in England were extracted from NHS Digital on 28 February 2023. Weekly total numbers of new cases per trust were calculated. COVID-19-related hospitalisations and deaths data in England between October 2020 and March 2022 were extracted from ONS. We used these data to calculate hospitalisation fatality ratios (HFRs) for our economic analysis. More details about the data accession processes are provided in the S1 Appendix Methods, 1.1–1.4.

### Statistical analysis

#### Analysis timeline

All analyses were conducted for four time periods to account for changes in healthcare testing policies, the vaccine rollout and the availability of data (Table 1). The four time periods were November 2020 to December 2020, January 2021 to July 2021, August 2021 to November 2021, and December 2021 to March 2022.

#### Association between testing and HCW absenteeism

The primary outcome for our study was FTE days lost due to COVID-19, expressed as a proportion of total FTE days. This was calculated for each acute trust and month of the study, as only monthly absenteeism data were available. The primary exposure variable was LFD test coverage, calculated as the number of tests reported per HCW, for each trust and month. (For further details see S1 Appendix – Methods, 1.5.) We used a mixed-effects linear model to examine the association between FTE days lost and LFD test coverage. The model was adjusted for community prevalence of COVID-19, average income deprivation score, relative prevalence of Alpha/Delta/Omicron variants (on a scale of 0 to 1) in England and acute trust LFD positivity rate (defined as the number of positive LFDs per headcount, calculated for each trust and month). Random intercepts for trust and month were included to account for data clustering. Vaccination data were not included in the final model, due to the sparsity of relevant data and low variability over time.

#### Association between testing and nosocomial infections

The secondary outcome of our investigation was the prevalence of nosocomial COVID-19 infections in NHS acute trusts, as reported to the ISARIC database. (For further details see S1 Appendix – Methods, 1.6.) The relationship between weekly prevalence of nosocomial infections and LFD test coverage in the previous week was modelled using logistic regressions weighted by the number of new COVID-19 cases reported in the ISARIC dataset for that week. Week and trust were fitted as fixed effects (due to identifiability issues in fitting random effects). The model was adjusted for the total number of new COVID-19 cases reported per trust/week. It was also adjusted for possible confounders: community prevalence, average income deprivation score, relative prevalence of Alpha/Delta/Omicron variants (on a scale of 0 to 1) and LFD positivity rate per trust-month in the previous week (defined as the number of positive LFD tests for a given trust and month/headcount for a given trust and month × 1000).

The model structures were compared by including cluster covariates (trust, month or week) and nonlinear forms of covariates and interaction terms with transmission, and the final models were selected based on the Akaike information criterion (AIC) and residuals. These models were then used to predict FTE days lost due to COVID-19 and the number of nosocomial infections in counterfactual scenarios where the LFD test intensity was set at 50%, 75%, 125% and 150% of the observed test level.

#### Economic analysis

The primary outputs of our economic analyses were the quality-adjusted life-years (QALYs) gained due to nosocomial infections averted within NHS acute trusts compared with various counterfactual LFD testing scenarios (50%, 75%, 125%, 150%, and 200% of the actual testing volume). The evaluation was conducted from the healthcare provider’s perspective, as the UK government funded all direct costs of testing and treatment for COVID-19. The time horizon of the analysis (October 2020 to March 2022) was determined by the period of interest for the UKHSA-commissioned retrospective independent evaluation of COVID-19 testing in England [13]. The secondary outputs of our economic analyses were the cost savings per nosocomial infection averted, per death averted, and QALYs gained within NHS acute trusts under various counterfactual LFD testing scenarios. We also conducted a sub-analysis for the two financial years (FY) in England covered by the study period, i.e. FY21, comprising the six-month period from October 2020 to March 2021; and FY22, comprising the full financial year from April 2021 to March 2022.

We present here the incremental costs per nosocomial infection averted, incremental costs per death averted and incremental cost-effectiveness ratio (ICER) (defined as the incremental cost per QALY gained) due to testing. Given the short time-horizon of the study, no discounting or adjustment for inflation was carried out. All costs and results were collected and are presented in pounds sterling (GBP). (For further details see S1 Appendix – Methods, 1.7.)

## Results

### HCW absenteeism

Monthly FTE days lost due to COVID-19 varied over time, by trust type and by staff group, ranging between 0% and 4% of the total corresponding FTE days available (Fig 1A). Monthly FTE days lost due to COVID-19 accounted for 0% to 60% of the total monthly FTE days lost for any reason (Fig S1). FTE days lost peaked at three timepoints: April 2020, January 2021 and January 2022, at 6.1%, 5.7% and 6.7%, respectively, with corresponding relative increases of 1.7-fold (90% confidence interval (CI) 1.5, 1.9), 1.2-fold (90% CI 1.2, 1.3) and 1.4-fold (90% CI 1.3, 1.5), respectively, compared with the median level from 2009 to 2019 (Fig 1B).

**Fig 1.**
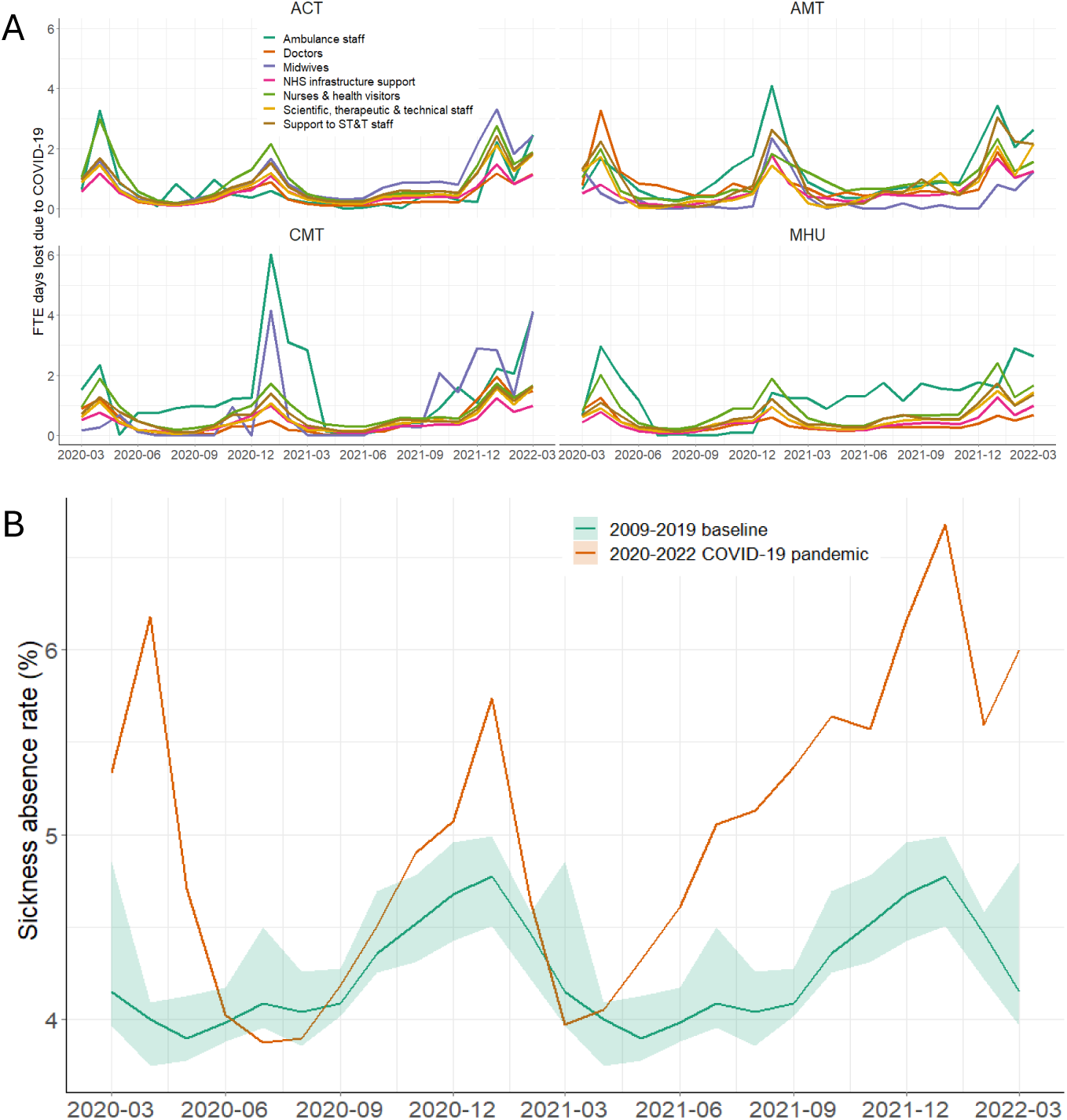
**Monthly absenteeism in NHS trusts in England during the COVID-19 pandemic from March 2020 to March 2022. (A) The percentage of monthly full-time equivalent (FTE) days lost due to COVID-19 by trust type and staff group over time (= monthly FTE days lost due to COVID-19 / monthly FTE days available × 100%). (B) Sickness absence rates in all staff groups at NHS acute trusts (= number of days off sick as a percentage of total workforce days contracted).** The orange line represents the overall percentage of FTE days lost; the green line with the shaded area indicates the median and 90% range of sickness absence rates from 2009 to 2019. ACT, acute trust; AMT, ambulance trust; CMT, community trust; MHU, mental health trust.

NHS staff absences due to sickness varied geographically, reflecting the sweep of the pandemic across England. In London and the southeast, absence rates increased more rapidly and to a higher peak than in other regions. Staff groups were also impacted differently by absences related to COVID-19, with doctors hit particularly hard in the first and third epidemic waves in England, in April 2020 and January 2022, respectively, when more than 40% of all doctors’ absences were due to COVID-19 (Fig S2). During the winter wave of January 2021, ambulance staff experienced the greatest number of FTE days lost due to COVID-19 (Fig S3). In acute trusts, variability between trusts was small and could possibly be explained by differences in staff composition (Table S1).

### Reported LFD test volumes, coverage and costs

Overall monthly volumes of reported LFDs followed similar trends across all healthcare settings except for community trusts and varied among the four time periods (Fig 2). A total of 16,352,554 LFDs were reported from October 2020 to March 2022, across the 136 acute trusts. A median of 0.83 (interquartile range, IQR 0.16–1.50) LFDs per HCW per week was reported between November and December 2020; 1.21 (IQR, 0.87–2.17) between January and July 2021; 0.71 (IQR, 0.62–0.79) between August and November 2021; and 0.77 (IQR, 0.60–1.2) between December 2021 and March 2022. The level of reporting also varied among trusts across the four evaluation periods, with the second time period (January 2021 to July 2021) having the highest average coverage (Table S2). Furthermore, 7 out of 136 trusts (5.0%) presented with less than one test per month for at least one month (Fig S4).

**Fig 2.**
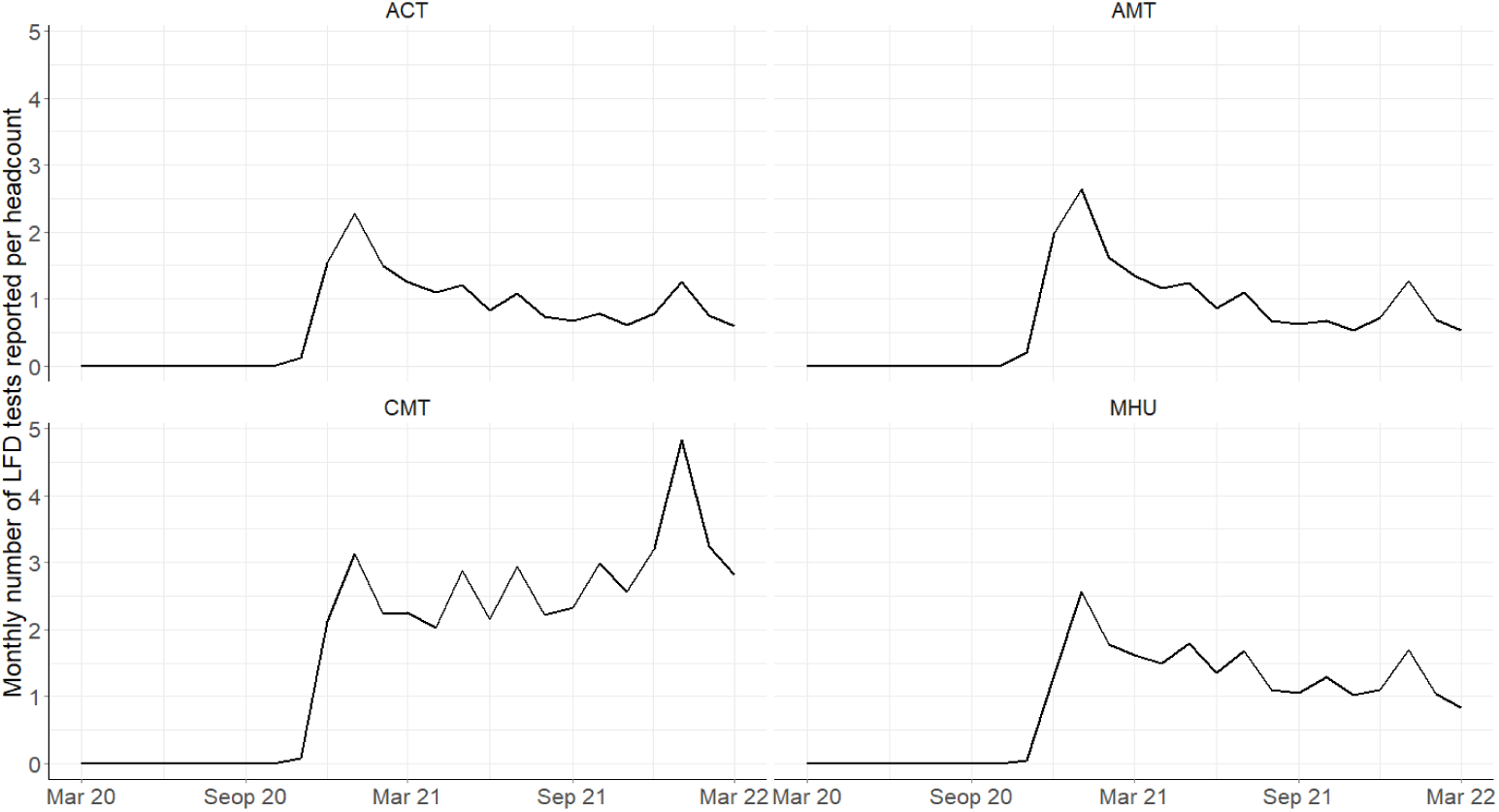
Monthly number^1^ of LFDs reported per HCW by trust type from March 2020 to March 2022^1^. ACT, acute trust; AMT, ambulance trust; CMT, community trust; MHU, mental health trust. ^1^LFDs were reported in MHU trusts in England, with a median 1.0 (interquartile range 0–2.0) tests per HCW per week; 0.68 (0.081–1.29) in November–December 2020; 1.69 (1.37–2.45) in January–July 2021; 1.08 (1.03–1.28) in August 2021–November 2021; and 1.07 (0.85– 1.65) in December 2021–March 2022. LFDs were reported in AMT trusts in England, with a median 0.58 (0–2.2) tests per HCW per week; 1.09 (0.24–1.93) in November–December 2020; 1.25 (0.90–2.50) in January–July 2021; 0.65 (0.53–0.68) in August 2021–November 2021; and 0.71 (0.55–1.23) in December 2021–March 2022. LFDs were reported in CMT trusts in England, with a median 2.1 (0–3.8) tests per HCW per week; 1.10 (0.13–2.07) in November–December 2020; 2.25 (2.05–3.09) in January–July 2021; 2.45 (2.22–2.96) in August 2021–November 2021; and 3.23 (2.85–4.71) in December 2021–March 2022.

The total financial cost of the healthcare testing service for the full evaluation period was GBP 1.77 billion, representing 7.6% of the total testing expenditure in England. This cost did not include payments made to HCWs for isolating following a contact with a positive COVID-19 case. LFD costs comprised approximately 92.5% (GBP 1.64 billion) of the total costs of the HCW testing service. Approximately 55% of these costs were direct costs, 19% were indirect costs and the remainder were overhead costs. Overhead and indirect costs were marginally higher in FY21 than in FY22 due to the initial costs of setting up the testing service. The average unit cost of an LFD distributed under the healthcare testing service was GBP 11.68. Overhead costs were only attributed to Pillar 2 testing.

### Association between FTE days lost and test coverage

Associations between the proportion of FTE days lost due to COVID-19 and test coverage, as well as other covariates, are shown in Fig S5–S10. No apparent associations between test coverage and percentage FTE days lost due to COVID-19 were observed except during the time period January 2021 to July 2021 (Fig S11). The increase in LFD test coverage was associated with decreases in FTE days lost due to COVID-19 during the first two time periods (Table 2, models 1 and 2). During the next two periods (Table 2, models 3 and 4), no such association was observed. Higher community prevalence levels were associated with significant increases in FTE days lost due to COVID-19 in all periods except for the pre-vaccination period. Effect sizes ranged from 0.23% to 0.46% increases in FTE days lost for each 1% relative increase in the community prevalence of COVID-19. Similarly, LFD positivity rates in HCWs were positively associated with FTE days lost due to COVID-19. Average income deprivation scores were not associated with lost FTE days. The changes in testing levels (50– 150%) were estimated to result in modest changes in FTE days lost due to COVID-19 for all time periods (Fig 3A).

**Fig 3.**
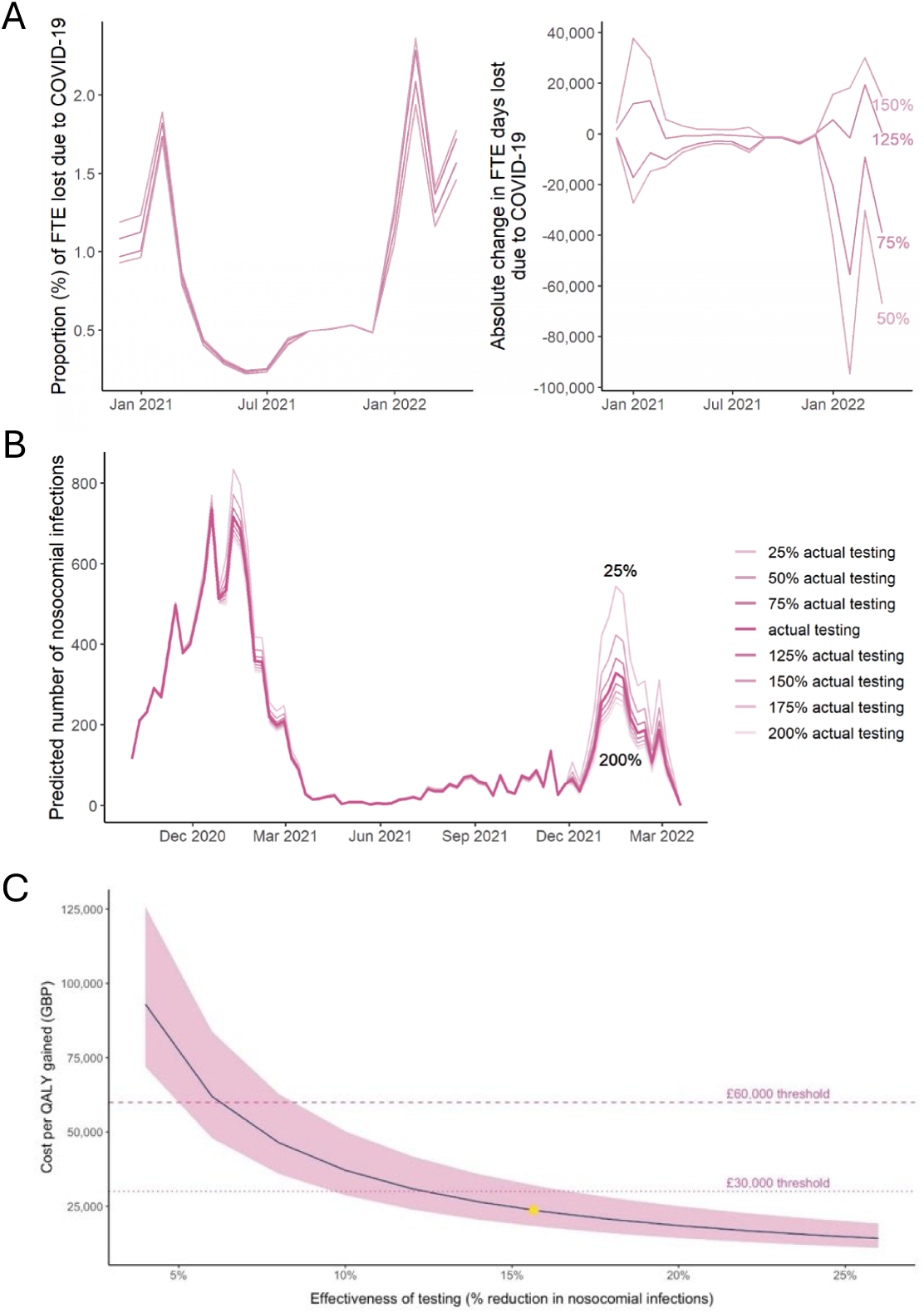
**(A) Predicted proportion and absolute FTE days lost due to COVID-19 in all acute trusts. (B) Weekly number of nosocomial infections predicted for different testing scenarios, for 136 trusts included in the analysis. (C) Cost effectiveness of weekly testing at different levels of testing effectiveness with respect to averting nosocomial COVID-19 infections in England.** The shaded area indicates the cost per QALY gained at an upper value of 8.8 QALYs per death averted and a lower value of 4.98 QALYs per death averted. The line shows the analysis conducted at a value of 6.78 QALYs per death averted. The yellow point at 16.8% represents the statistically modelled level of testing effectiveness in reducing nosocomial infections.

**Table 2.**
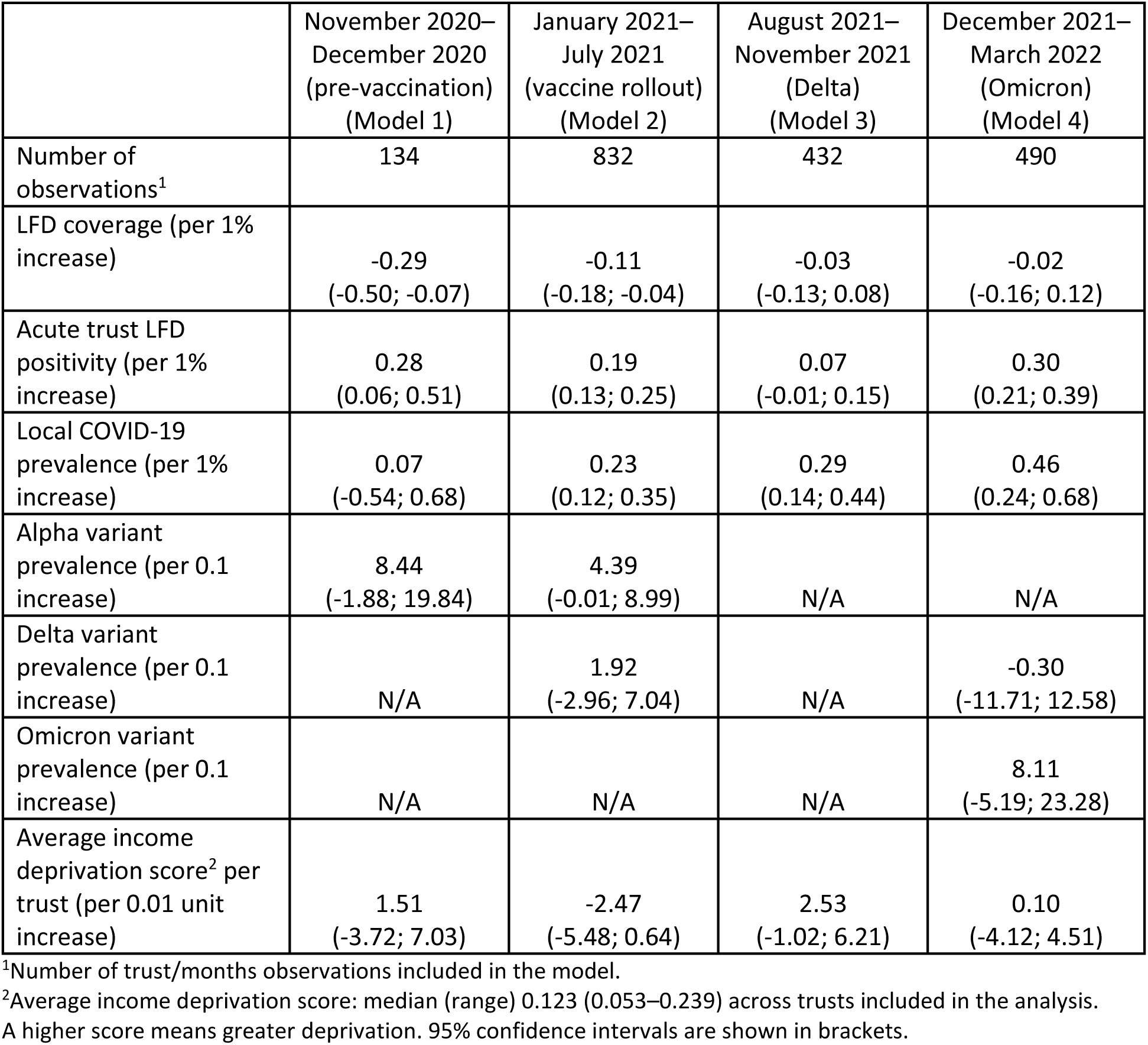
Estimated relative change (%, with 95% CI) in the proportion of the total available FTE days lost due to COVID-19 in the four time periods examined. These estimates are from linear regression models as described in the text.

### Nosocomial infections in NHS acute trusts

In the ISARIC database, 136 NHS acute trusts were represented, with data available for a median of 54 (range 1–75) weeks. Overall, 3794 nosocomial infections after 7 days of admission (2049 after 14 days of admission, Table S3) were identified among 106,377 hospitalised COVID-19 patients (Table S4). The median (range) number of new COVID-19 cases per week was 9 (1–531) with a median (range) of 0 (0–40) nosocomial infections identified using 7 days as the cut off. Table 3 shows the distribution of trust-level parameters for hospitalised COVID-19 patients with/without nosocomial infections.

**Table 3.** Summary statistics for measured covariates in hospitalised COVID-19 cases, by type of COVID-19 infection.

| Parameter | Nosocomial infection | Summary statistics |  |  |  |  |
| --- | --- | --- | --- | --- | --- | --- |
|  |  | Minimum | p25* | Median | p75* | Maximum |
| November–December 2020 |  |  |  |  |  |  |
| LFD coverage (per 1000) | No | 0 | 0 | 0.052 | 68.7 | 2121 |
|  | Yes | 0 | 0 | 0 | 84.3 | 2121 |
| Acute trust LFD positivity (per 1000) | No | 0 | 0 | 0 | 1.37 | 64.4 |
|  | Yes | 0 | 0 | 0 | 1.45 | 64.4 |
| Local COVID-19 prevalence | No | 0.001 | 0.008 | 0.011 | 0.016 | 0.031 |
|  | Yes | 0.001 | 0.007 | 0.010 | 0.014 | 0.030 |
| Alpha variant prevalence | No | 0 | 0 | 0.008 | 0.215 | 1 |
|  | Yes | 0 | 0 | 0.014 | 0.264 | 1 |
| Average income deprivation score | No | 0.053 | 0.115 | 0.141 | 0.167 | 0.240 |
|  | Yes | 0.053 | 0.108 | 0.126 | 0.154 | 0.235 |
| January–July 2021 |  |  |  |  |  |  |
| LFD coverage (per 1000) | No | 0 | 176 | 352 | 523 | 1848 |
|  | Yes | 0 | 221 | 417 | 618 | 1478 |
| Acute trust LFD positivity (per 1000) | No | 0 | 1.25 | 5.38 | 17.6 | 134 |
|  | Yes | 0 | 2.00 | 7.43 | 22.9 | 134 |
| Local COVID-19 prevalence | No | 0.0001 | 0.007 | 0.010 | 0.015 | 0.034 |
|  | Yes | 0.0001 | 0.006 | 0.010 | 0.015 | 0.029 |
| Alpha variant prevalence | No | 0 | 0.335 | 0.872 | 0.949 | 1 |
|  | Yes | 0 | 0.699 | 0.898 | 0.956 | 1 |
| Delta variant prevalence | No | 0 | 0 | 0 | 0.323 | 1 |
|  | Yes | 0 | 0 | 0 | 0 | 1 |
|  | No | 0 | 0 | 0 | 0 | 0.012 |
| Omicron variant prevalence | Yes | 0 | 0 | 0 | 0 | 0.011 |
| Average income deprivation score | No | 0.053 | 0.110 | 0.137 | 0.167 | 0.239 |
|  | Yes | 0.053 | 0.108 | 0.122 | 0.147 | 0.235 |
| August–November 2021 |  |  |  |  |  |  |
| LFD coverage (per 1000) | No | 0 | 58.8 | 127 | 225 | 1432 |
|  | Yes | 0 | 75.3 | 126 | 220 | 1305 |
| Acute trust LFD positivity (per 1000) | No | 0 | 0.739 | 2.01 | 3.53 | 289 |
|  | Yes | 0 | 0.858 | 2.22 | 3.58 | 14.6 |
| Local COVID-19 prevalence | No | 0.003 | 0.009 | 0.011 | 0.013 | 0.026 |
|  | Yes | 0.003 | 0.010 | 0.011 | 0.013 | 0.021 |
| Alpha variant prevalence | No | 0 | 0 | 0 | 0 | 0.004 |
|  | Yes | 0 | 0 | 0 | 0 | 0.0002 |
| Delta variant prevalence | No | 0.964 | 0.997 | 1 | 1 | 1 |
|  | Yes | 0.975 | 0.998 | 1 | 1 | 1 |
| Omicron variant prevalence | No | 0 | 0 | 0 | 0 | 0.013 |
|  | Yes | 0 | 0 | 0 | 0 | 0.008 |
| Average income deprivation score | No | 0.057 | 0.108 | 0.135 | 0.168 | 0.239 |
|  | Yes | 0.057 | 0.108 | 0.141 | 0.184 | 0.235 |
| December 2021–March 2022 |  |  |  |  |  |  |
| LFD coverage (per 1000) | No | 0 | 61.9 | 146 | 276 | 2437 |
|  | Yes | 0 | 85.1 | 158 | 276 | 1240 |
| Acute trust LFD positivity (per 1000) | No | 0 | 2.49 | 14.9 | 33.3 | 479 |
|  | Yes | 0 | 5.13 | 24.1 | 41.9 | 144 |
| Local COVID-19 prevalence | No | 0.006 | 0.023 | 0.033 | 0.041 | 0.057 |
|  | Yes | 0.008 | 0.028 | 0.035 | 0.039 | 0.051 |
| Delta variant prevalence | No | 0 | 0.001 | 0.015 | 0.199 | 1 |
|  | Yes | 0 | 0.0002 | 0.006 | 0.038 | 1 |
| Omicron variant prevalence | No | 0 | 0.768 | 0.962 | 0.981 | 1 |
|  | Yes | 0 | 0.936 | 0.976 | 0.985 | 1 |
| Average income deprivation score | No | 0.057 | 0.116 | 0.141 | 0.170 | 0.235 |
|  | Yes | 0.066 | 0.122 | 0.141 | 0.166 | 0.235 |

\*p25 = 25% centile; p75 = 75% centile.
LFD coverage, LFD positivity and prevalence data are per week.

### Association between nosocomial infections and test coverage

The proportion of nosocomial infections among new weekly cases in hospitalised patients (which included new admissions and cases diagnosed in hospital) was negatively associated with reported LFD testing levels (Fig 3B). However, the strength of the association varied over time and was estimated to be highest during the Omicron period, with a doubling of the test coverage associated with 22% (95% CI 4%, 37%) fewer nosocomial infections (Table 4). Our model predicted that the observed HCW testing/reporting was associated with a 16.8% (95% CI 8.2%, 18.8%) reduction in nosocomial infections compared with a hypothetical test scenario at 25% of actual levels (Fig 3B, Fig S11 and Table S5). A 0.1% increase in the prevalence of the Omicron variant, compared with circulating wild-type or Delta variants, was associated with a 35% increase in the odds of nosocomial infections among hospitalised COVID-19 cases (odds ratio (OR) = 1.35, 95% CI 1.07, 1.70, presented in Table 4 when changes in Omicron were compared with any other circulating variants). No association between nosocomial infections and LFD positivity rate in HCWs, or the population prevalence, was observed for any of the time periods. A negative association was observed between nosocomial infections and average income deprivation score, although this analysis was only conducted for the first two time periods, due to identifiability issues. Similar results were obtained in the sensitivity analysis when a nosocomial infection was defined as a patient who developed COVID-19 symptoms after 14 days since their admission (Table S6).

**Table 4.** Logistic regression model for the prevalence of nosocomial COVID-19 infections among all COVID-19 infections in hospitalised patients. Odds ratios (95% CI) are shown.

|  | November 2020–<br>December 2020<br>(pre-vaccination)<br>(1) | January 2021–<br>July 2021<br>(vaccine rollout)<br>(2) | August 2021–<br>November 2021<br>(Delta)<br>(3) | December<br>2021–March<br>2022 (Omicron)<br>(4) |
| --- | --- | --- | --- | --- |
| Number of<br>observations<sup>1</sup> | 1314 | 2852 | 1489 | 829 |
| LFD coverage (per<br>100% increase) | 0.98<br>(0.96, 0.99) | 0.92<br>(0.89, 0.95) | 1.06<br>(0.89, 1.26) | 0.78<br>(0.63, 0.96) |
| Acute trust LFD<br>positivity (per<br>1/100 increase) | 0.90<br>(0.82, 1.00) | 0.96<br>(0.89, 1.04) | 0.87<br>(0.53, 1.42) | 1.03<br>(0.97, 1.09) |
| Local COVID-19<br>prevalence (per<br>1/100 increase) | 0.90<br>(0.74, 1.09) | 0.76<br>(0.53, 1.11) | 0.84<br>(0.45, 1.53) | 1.02<br>(0.82, 1.27) |
| Alpha variant prevalence (per 0.1 increase) | 1.02<br>(0.96, 1.08) | 1.00<br>(0.92, 1.08) | N/A | N/A |
| Delta variant prevalence (per 0.1 increase) | N/A | 1.00 (0.80, 1.24) | Delta variant prevalent | 1.33<br>(0.67, 2.66) |
| Omicron variant prevalence (per 0.1 increase) | N/A | N/A | N/A | 1.72 (0.90, 3.27) |
| Average income deprivation score<sup>2</sup> per acute trust (per 0.01-unit increase) | 0.16<br>(0.07, 0.33) | 0.53<br>(0.27, 1.07) | N/A | N/A |

<sup>1</sup>Number of acute trust/week observations included in the model.
<sup>2</sup>Average income deprivation score, median (range) 0.130 (0.053–0.239) across trusts included in the analysis. A higher score means greater deprivation; 95% confidence intervals are shown in brackets. The results are adjusted for trust and calendar week (as fixed effects) and the number of new COVID-19 cases identified at the trust that week.
N/A, not applicable

### Economic analysis of LFD testing on nosocomial infections averted

At a testing effectiveness of 8% to 20%, between 17,500 and 43,800 nosocomial infections were averted during weekly testing of HCWs. The number of deaths averted ranged from 5500 to 13,800, with a saving per death averted of GBP 127,900–320,800. Between 38,100 and 95,100 QALYs were gained, translating to a value per QALY gained of GBP 18,500–46,400. A sensitivity analysis of the QALY values for deaths generated a saving of GBP 25,000–62,800 per QALY gained at a value of 4.98 QALYs per death averted and GBP 14,300–35,900 per QALY gained at a value of 8.8 QALYs per death averted at a testing effectiveness of 4% to 26% (Table 5).

**Table 5.** Summary of the cost effectiveness of weekly and daily testing with respect to nosocomial infections averted during the evaluation period (October 2020 to March 2022).

|  | Weekly testing |  |  | Daily testing |  |  |
| --- | --- | --- | --- | --- | --- | --- |
| Testing level | 8% | 16% | 20% | 25% | 15% | 30% |
| Number of nosocomial infections averted | 17,500 | 35,000 | 43,800 | 50,400 | 32,800 | 65,700 |
| Number of deaths averted due to nosocomial infections averted | 5,500 | 11,000 | 13,800 | 15,800 | 10,300 | 20,700 |
| Cost per nosocomial infection averted (GBP) | 101,100 | 50,600 | 40,5500 | 35,200 | 53,900 | 27,000 |
| Cost per death averted (GBP) | 320,800 | 160,000 | 127,900 | 410,800 | 630,300 | 314,800 |
| Number of QALYs gained | 38,000 | 76,100 | 95,100 | 109,400 | 71,400 | 142,700 |
| Cost per QALY gained (GBP) | 46,400 | 23,200 | 18,500 | 59,500 | 91,300 | 45,600 |

The statistical analysis presented estimated that HCW testing was associated with a 16.8% (95% CI 8.2%, 18.8%) reduction in nosocomial infections compared with a testing scenario at 25% of actual levels.

At a daily testing effectiveness of 15% to 30%, between 32,800 and 65,700 nosocomial infections were averted due to testing of HCWs. The number of deaths averted ranged from 10,300 to 20,700, with a cost per death averted of GBP 314,800–630,300. The cost per QALY gained was GBP 45,600–91,300. A sensitivity analysis of the QALY values per death averted found this would cost GBP 61,600–123,400 per QALY gained at a value of 4.98 QALYs per death averted and GBP 35,300–70,600 per QALY gained at a value of 8.8 QALYs per death averted (Table 5).

The ICER (incremental costs per QALY gained) for various assumptions of testing effectiveness with respect to reducing nosocomial COVID-19 infections in acute trusts in England are illustrated in Fig 3C. The range (shaded area) shown in the plot indicates the cost per QALY gained at an upper value of 8.8 QALYs per death averted and a lower value of 4.98 QALYs per death averted. The HCW testing service was estimated to be cost effective at reducing nosocomial infections at values of effectiveness of 5.5% or more, at a willingness to pay threshold of GBP 70,000. Our statistical analysis estimated that HCW testing was associated with a 16.8% (95% CI 8.2%, 18.8%) reduction in nosocomial infections compared with a testing scenario at 25% of actual levels. This suggests that for all values of effectiveness predicted in the 95% CI, the HCW testing service was cost effective.

The yellow dot in Fig 3C indicates that at an overall 16.8% decrease in nosocomial infections, the HCW testing service was cost effective, at a cost of GBP 22,100 per QALY gained. This is well below the willingness to pay threshold of GBP 70,000 published by the UK government [20]. The HCW testing service was also cost effective at reducing nosocomial infections at values of effectiveness of 12.5% or more at the National Institute for Health and Care Excellence (NICE) threshold of GBP 30,000 [21]. Compared with a hypothetical scenario of no testing, the HCW testing service contributed to an even greater reduction in nosocomial infections and hence was highly cost effective.

## Discussion

During the COVID-19 pandemic, in the UK and worldwide, HCWs suffered from consistently high rates of exposure [22], a lack of personal protective equipment (PPE) [23] and high levels of mental anxiety [24], among other stressors. The UK’s NHS subsequently experienced major disruption due to staff absences and the burden of nosocomial infections [25, 26]. A better understanding of the burden of staff absenteeism and nosocomial infections among HCWs due to COVID-19 and the practical impact of large-scale LFD testing on reducing this burden will be integral to future pandemic preparedness plans in healthcare settings.

We have presented a timeline of FTE days lost among NHS staff due to COVID-19, from October 2020 to March 2022, which accounted for more than 50% of the total monthly FTE days lost for any reason, an average 1.5-fold increase during the first peak of the COVID-19 epidemic in March 2020 compared with the 2009–2019 median level. This situation worsened across certain trusts during the Omicron phase in February 2022. There was substantial heterogeneity in FTE days lost by geographical locations of trusts, trust type and staff group, likely reflecting the heterogeneity of COVID-19 transmission and the risks associated with being an HCW [27]. Among 136 NHS acute trusts, we identified that nosocomial infections accounted for 2.9% to 3.5% of the total 106,377 hospitalised COVID-19 patients from October 2020 to March 2022. These estimates are around 1% higher than a previous estimate of nosocomial infections in England between June 2020 and March 2021 [28] but dramatically lower than reported from other studies that focused on the early days of the pandemic [3, 29–31]. The reported number of LFD tests from NHS acute trusts was likely to be an underestimate of the actual number of tests performed. Possible reasons for this discrepancy include biases in reporting, e.g. some negative results may not have been reported as there was no infection prevention and control (IPC) action required for the individual HCW themselves or for their trust; furthermore, some individuals may not have reported positive results if there would have been an impact on their personal income or the income of their family due to them having to isolate.

Testing could potentially have two different effects on absenteeism, which work in opposite directions, where nosocomial infection is also an important mediator. Testing meant more cases would be identified and would need to isolate (both necessary isolation of asymptomatic cases and isolation of individuals who received false-positive results), leading to an increase in absenteeism. On the other hand, the isolation of individuals with true-positive results would reduce onward transmission and subsequent isolations, leading to a reduction in staff absenteeism. The balance of these two dynamics might change over time and at different comparator levels of testing. Our modelling results imply that increasing the volume of asymptomatic testing prior to August 2021 reduced absence rates among HCWs, but this impact was subsequently diminished by the high level of vaccination coverage. Similar observations have been reported elsewhere. For example, a hospital in the Canadian city of Campinas experienced a significantly reduced level of hospital absenteeism due to COVID-19 from March 2020 to September 2020 by using PCR tests for screening and then requiring the isolation of asymptomatic HCWs who had tested positive, resulting in a reduction in transmission of COVID-19 to hospitalised patients [32]. Using a computational model of SARS-

CoV-2 transmission in a hospital in England, another study estimated that testing asymptomatic patients on admission would reduce the rate of nosocomial SARS-CoV-2 infection by 8.1% to 21.5% [33]. On the other hand, another modelling study showed that, compared with no testing, testing all HCWs would prevent most transmission, but at the expense of greatly increased staff absences and greater capacity for testing needed [34]. That study found that the optimal testing strategy to reduce staff absences would be to test HCWs in household quarantine if they had been exposed to symptomatic household contacts. This would increase the risk of workplace transmission; however, the authors noted that this risk could be mitigated, although not eliminated, by re-testing initially negative samples of staff who were quarantining [34].

Our model suggests that high levels of vaccination coverage might dilute the effect of testing on reducing absenteeism, which could be attributed to the vaccine’s efficacy in blocking infections/transmissions and resultant changes in isolation policies (after vaccination the isolation period following a positive test result could be shortened) [35]. An observational study involving a prospective cohort of 4964 HCWs in Canada showed that vaccination was strongly related to shorter absences due to COVID-19-related illness [36].

Although the rollout of vaccination may have made ongoing testing appear to be marginally less cost effective, continued testing should be considered a reasonable and overall cost-effective insurance strategy given the uncertainties around the long-term effectiveness of vaccines and the potential for future VOCs. This also raises the question of whether a standard health economics approach is appropriate for analyses such as this, post-vaccination rollout. In England, testing was continued based on the precautionary principle and because the potential impact of new variants was unknown. Traditional cost-effectiveness analyses do not take into account the willingness to pay for this type of assurance. We used a provider approach for our economic analysis. A societal approach that incorporates the economic cost of illness, including losses due to absenteeism, while not accounting for the assurance against uncertainty, is likely to result in testing being more cost effective even after vaccination. At the same time, an analysis conducted for different time periods may have shown that the intervention was not cost effective, especially with VOCs that were less severe or transmissible, e.g. Omicron.

Higher community prevalence levels were associated with significant HCW absenteeism even after adjusting for testing levels. This suggested that although testing could have a positive impact on mitigating outbreaks within healthcare settings and reducing HCWs’ absenteeism at a given level of community transmission rate, the overall community prevalence placed greater external pressure on outbreak management and HCW attendance. We might anticipate HCW absence rates to be higher when there is greater community transmission, but if testing is particularly protective at that point, then testing would play a key role in outbreak mitigation within healthcare settings.

We estimated LFD test coverage was negatively correlated with nosocomial infections, although the strength of this correlation varied over time and was estimated to be highest during the Omicron period. These findings are consistent with mechanistic modelling results, which estimated nosocomial incidence could be reduced by up to 40–47% (range of means) with routine symptomatic testing using reverse transcriptase PCR (RT-PCR), 59–63% with the addition of a timely round of antigen rapid diagnostic testing screening and 69–75% with well-timed two-round screening [37]. The risk of developing nosocomial infections was estimated to vary for different VOCs. Another modelling study demonstrated that the risk of a highly transmissible (including pre-symptomatic transmission) variant could amplify nosocomial infections, e.g. it was estimated a variant with 56% higher transmissibility would increase nosocomial transmissions by 303% [38]. Our model predicted that the observed HCW testing/reporting was associated with a 16.8% (95% CI 8.2%, 18.8%) reduction in nosocomial infections compared with a hypothetical testing scenario at 25% of actual levels. This was in line with the findings from an internal modelling exercise carried out by UKHSA in 2022, which estimated that the reductions in nosocomial infections due to weekly and daily testing were 16% and 25.4%, respectively. Combined with testing and treatment costs, our cost-effectiveness analysis estimated that the HCW testing service was cost effective at reducing nosocomial infections when effectiveness attained 5.5% or more, at a willingness to pay threshold of GBP 70,000.

Our study has some strengths and limitations. To the best of our knowledge, this is the first study to examine the association between test coverage and nosocomial COVID-19 transmission in England, incorporating an economic analysis. The data on 16 million LFDs represent a uniquely large source of information, with high coverage of the HCW population who could access tests for free during the pandemic. However, data were not available that would have enabled us to quantify the impact of patients with undetected COVID-19 being released into the community and potentially infecting others through secondary transmission. We could also not account for the effects of long COVID-19 in our economic analysis, as these effects could not be quantified. Furthermore, the nosocomial infections estimated using the ISARIC dataset are likely to be biased by the inclusion criteria, in that only individuals who consented to participate in the study were included.

Our study has implications for the development of testing policies in the healthcare setting for future pandemics, particularly for respiratory pathogens similar to COVID-19. Our evaluation also showed that there was a discrepancy between the numbers of LFD tests distributed and reported in NHS trusts, likely suggesting a gap between testing and test reporting, a general issue across various settings [39]. Our analysis of the available data indicated that HCW testing interventions had varying impacts (on both nosocomial infections and HCW FTE days lost) throughout the pandemic, possibly influenced by external factors such as community prevalence and vaccination. This finding highlights the importance and necessity of developing more targeted and agile testing systems, which operationally would require the ability to turn mass testing off and on as an epidemic progress and was described as the primary intention of the retrospective pan-evaluation of the mass testing conducted during the COVID-19 pandemic [13]. The rollout of HCW testing interventions through pilot studies, with collection of and/or timely access to data relating to suitable endpoints (including HCW absenteeism, routine test results, community prevalence, and hospitalisation and mortality data), could be used by relevant authorities to support the real-time assessment of any testing service and adjustment of testing interventions.

## EY-Oxford Health Analytics Consortium membership list

Ricardo Aguas, Ma’ayan Amswych, Billie Andersen-Waine, Sumali Bajaj, Kweku Bimpong, Adam Bodley, Liberty Cantrell, Siyu Chen, Richard Creswell, Prabin Dahal, Sophie Dickinson, Sabine Dittrich, Tracy Evans, Angus Ferguson-Lewis, Caroline Franco, Bo Gao, Rachel Hounsell, Muhammad Kasim, Claire Keene, Ben Lambert, Umar Mahmood, Melinda Mills, Ainura Moldokmatova, Sassy Molyneux, Reshania Naidoo, Randolph Ngwafor Anye, Jared Norman, Wirichada Pan-Ngum, Sarah Pinto-Duschinsky, Sunil Pokharel, Anastasiia Polner, Katarzyna Przybylska, Emily Rowe, Sompob Saralamba, Rima Shretta, Sheetal Silal, Kasia Stepniewska, Joseph Tsui, Merryn Voysey, Marta Wanat, Lisa J. White, Gulsen Yenidogan

## Data Availability

The data supporting the findings of this study are available within the paper and its supporting information. The data were made available by UKHSA to the manuscript's authors as part of a retrospective evaluation of England's COVID-19 testing programme. The authors cannot make the underlying datasets publicly available for ethical and legal reasons, particularly given the sensitive nature of the information included. Applications for access to the anonymised data should be submitted to UKHSA.

## Author Contributions

LJW, KS and RS conceived and designed the study. LJW, MV, RS, and KS supervised the work. SC, RH, LC, LHT, KS and RS coded the models and analysed the data. SC and RN collected the testing policy data. KS, MV, LJW, and RS advised on the methodologies. SC, RH, LC, RS and KS wrote the initial draft of the manuscript. All authors contributed to guiding the research questions, interpretation of results, and reviewing and approving the manuscript. The data were restricted due to privacy concerns and only available to a subset of the authors. The corresponding and lead authors had full access to all data used in the study.

## Ethics approval

The study protocol for the evaluation project, which this research fed into, was granted ethics approval by the UKHSA Research Ethics and Governance Group, reference number NR0347. All relevant ethics guidelines were followed throughout.

## Acknowledgements

Medical editing support in the preparation of this paper was provided by Adam Bodley, according to Good Publication Practice. The authors would like to express their gratitude to Oliver Munn, Sarah Tunkel, Nick Sharp, Sariyu Shoge and Olutoye Olatunbosun of UKHSA for sponsoring this research and enabling access to the data used in this study.

## Abbreviations

ACT: acute trust
AIC: Akaike information criterion
AMT: ambulance trust
CI: confidence interval
CMT: community trust
COVID-19: coronavirus disease 2019
FY: financial year
HCHS: Hospital and Community Health Service
HCW: healthcare worker
ICER: incremental cost-effectiveness ratio
IQR: interquartile range
ISARIC: International Severe Acute Respiratory and emerging Infection Consortium
LFD: lateral flow device
LTLA: lower-tier local authority
MHU: mental health trust
NHS: National Health Service
NIMS: National Immunisation Management System
ONS: Office for National Statistics
OR: odds ratio
PCR: polymerase chain reaction
QALY: quality-adjusted life-year
SARS-CoV-2: severe acute respiratory syndrome coronavirus 2

